# Distinct transcriptomic patterns in bicuspid aortic valve aortopathy suggest a unique mechanism of ascending aortic aneurysm progression

**DOI:** 10.64898/2026.05.19.26353631

**Authors:** Lauren E. Levy, John Chamberlin, Andrea M. Steely, Vikas Sharma, Matthew L. Goodwin, Hiroshi Kagawa, Michael Seipp, Sara Pereira, Craig H. Selzman, Aaron Quinlan, Martin Tristani-Firouzi, Jason P. Glotzbach

**Affiliations:** Division of Cardiothoracic Surgery, Department of Surgery, University of Utah, Salt Lake City, Utah, USA; Department of Human Genetics, University of Utah, Salt Lake City, Utah, USA; Department of Surgery Research Core Lab, University of Utah, Salt Lake City, Utah, USA; Department of Pediatrics, Division of Cardiology, University of Utah, Salt Lake City, Utah, USA

## Abstract

**Objective:** To compare RNA-sequencing-derived transcriptomic profiles of thoracic aortic aneurysm tissue from individuals with bicuspid versus trileaflet aortic valves.

**Methods:** Human ascending aortic tissue was collected from patients undergoing cardiac surgery at a single institution between January 2021 and December 2022 with bicuspid aortic valves (BAV) and trileaflet aortic valves (TAV) with (-A) and without (-N) thoracic aortic aneurysm. TAV-N tissue was collected from heart transplant donors. The decision to perform ascending aortic replacement was at surgeon discretion following ACC/AHA guidelines. Bulk RNA was extracted from the aortic wall, and Illumina RNA Sequencing performed. Differential gene expression analysis, enrichment analyses, network analysis, and deconvolution single cell-mapping were performed in R. Cell-type specificity of differentially expressed genes was determined using an established Aorta single cell RNA sequencing matrix.

**Results:** Tissue samples from 60 patients were included: 4 TAV-N, 16 BAV-N, 28 BAV-A, and 12 TAV-A. Average absolute aortic diameter was 5.1 ± 0.38 cm for BAV-A and 5.3 ± 0.44 cm for TAV-A, as measured on pre-operative CT. Gene ontology analyses of differentially expressed genes revealed enrichment of genes associated with extracellular matrix (ECM) organization, cellular receptor interactions and vascular smooth muscle cell (VSMC) function in BAV-A and BAV-N. In contrast, analysis of TAV-A versus TAV-N showed enrichment in genes associated with immune and inflammatory processes. Cell-type specificity analysis revealed a downregulation of genes associated with ECM components, cell signaling, and ECM remodeling in mesenchymal cells, VSMCs, and matrix fibroblasts specifically in BAV-A versus BAV-N.

**Conclusions:** The transcriptome changes observed in aneurysmal aortas of BAV and TAV patients are distinct, suggesting mechanistic differences contributing to aneurysm development and progression. The observed differences in gene expression between the non-aneurysmal aortas may signify a predisposition to aneurysm development unique to BAV aortopathy.

## Introduction

Bicuspid aortic valve (BAV) occurs when two of the three aortic valve cusps are fused during heart development. This condition is the most common congenital heart defect, occurring in 1-2% of the Western population^1–6^. Up to 50% of BAV patients will develop an associated aortopathy (thoracic aortic aneurysm (TAA) or dissection), exposing 1.5-3 million people in the U.S. to significant morbidity and mortality^7^. Type A aortic dissection (involving the proximal thoracic aorta) is a deadly emergency, with a 50% out-of-hospital and 0.5% per hour mortality rate. Patients who survive transport to a hospital and undergo emergency surgery suffer an operative mortality of 15-30% ^8–11^. Conversely, operative mortality is 1-3% for elective thoracic aortic replacement for aneurysm, a procedure that virtually eliminates the risk of mortality from subsequent Type A aortic dissection^12^. However, not all BAV patients will develop an aortic dissection or even an ascending aortic aneurysm, so identifying which patients are at risk and should undergo elective repair—and when—is critical. Current AHA/ACC clinical guidelines are imaging-based, defining an aortic diameter ≥ 5.5 cm and/or growth rate ≥ 0.5 cm/year as Class I indications for elective ascending aortic surgery^13^, yet up to 50% of patients who suffer acute aortic dissection have an aortic size below the threshold for elective aortic replacement^13,14^. The current size-based strategy to predict aortic dissection risk and guide management of BAV aortopathy is inadequate as a risk stratification tool.

BAV and BAV aortopathy typically lack other external characteristic clinical features and are often clinically silent before being diagnosed incidentally or at the time of aortic dissection. However, unlike other non-syndromic thoracic aortopathies, BAV aortopathy is thought to have a heritable component with incomplete penetrance and variable expressivity^15,16^. Previously, we demonstrated a significant familial association with BAV aortopathy and an increased risk of aortic dissection in relatives of patients with BAV aortopathy compared to the general population^17^. These results suggest that BAV aortopathy may have a genomic predisposition^18^. Variants in several genes have been associated with BAV aortopathy and non-syndromic thoracic aortic aneurysms, including *NOTCH1*^19–21^, *TGFBR2*^22^, *ACTA2*^23^, *FBN1*^24,25^, and *MYH11*^26–29^. Mutations in these genes result in inappropriate ECM remodeling, VSMC dysfunction, and TGF-β signaling dysregulation^30^. However, many of these studies are underpowered for BAV or do not distinguish between BAV and trileaflet aortic valves (TAV) in their TAA patients.

Novel variant discovery is challenging in bicuspid aortopathy due to the phenotypic heterogeneity observed across the population as well as the incomplete penetrance^31^. A recent large genome-wide association study meta-analysis identified 32 novel loci that are associated with bicuspid aortic valve^32^. This study did not focus on patients with aneurysm or dissection, and the genomic and mechanistic causes of BAV aortopathy remain elusive^33^. In order to understand the functional impact of associated genome variants, an understanding of the transcriptome-level changes in BAV aortopathy aortic tissue will be crucial^34^.

The purpose of this study was to test the hypothesis that BAV aortopathy progresses through a distinct molecular mechanism as compared to trileaflet aortic valve aneurysms using bulk RNA-sequencing.

## Methods

Ascending aortic tissue was collected from patients who consented to research undergoing cardiac surgery at the University of Utah between January 2021 and December 2022 with BAV and TAV with (-A) and without (-N) thoracic aortic aneurysm under IRB #00121538. Informed consent was obtained for all participants. Patients were screened prior to surgery based on aortic valve morphology (trileaflet versus bicuspid) on echocardiography and thoracic aortic size and subsequently classified into one of four distinct phenotype groups. TAV-N tissue was collected from heart transplant donors. Ascending aortic replacement was performed at surgeon discretion following ACC/AHA guidelines. Tissue was collected intra-operatively and immediately placed in RNAlater then stored at −80°C.

Bulk RNA was extracted from the aortic wall, and Illumina RNA Sequencing performed at the University of Utah HCI High-Throughput Genomics Core using previously established protocols. Briefly, RNA isolation was performed using the RNeasy Fibrous Tissue Mini Kit (Qiagen). Total RNA samples (5-500 ng) were hybridized with NEBNext rRNA Depletion Kit v2 (Human, Mouse, Rat) (E7400) to substantially diminish rRNA from the samples. Stranded RNA sequencing libraries were prepared as described using the NEBNext Ultra II Directional RNA Library Prep Kit for Illumina (E7760L). RNA concentration was measured with a Qubit RNA HS Assay Kit (Fisher Scientific #Q32855). RNA quality was evaluated with an Agilent Technologies RNA ScreenTape Assay (5067-5579 and 5067-5580). The molarity of adapter-modified molecules was defined by quantitative PCR using the Kapa Biosystems Kapa Library Quant Kit (cat#KK4824). Individual libraries were normalized and pooled in preparation for Illumina sequence analysis. Sequencing libraries were chemically denatured and applied to an Illumina NovaSeq flow cell using the NovaSeq XP workflow (20043131). Following transfer of the flowcell to an Illumina NovaSeq 6000 instrument, a 151 x 151 cycle paired end sequence run was performed using a NovaSeq 6000 S4 reagent Kit v1.5 (20028312). A single base mismatch protocol was used.

Demographics were obtained by review of the electronic medical record. Family history was defined as a known family member with thoracic aortic aneurysm or aortic dissection; family history of BAV was not included in this distinction. Absolute aortic diameter was measured on the patient’s most recent pre-operative Computed Tomography (CT) scan using three-dimensional reconstruction software. Valve morphology as well as presence and severity of aortic stenosis was defined from operative report and intraoperative pre-bypass transesophageal echocardiography (TEE) performed and read by a cardiac anesthesiologist.

Patients were excluded from bioinformatics analysis if they had tissue collected at the time of repair of thoracic aortic dissection or a heritable syndromic aortopathy (Marfan syndrome, Loeys-Dietz syndrome, autosomal dominant *ACTA2* mutation). Patients were additionally excluded if their RNA-Seq data failed quality control for RNA-Seq performed in R due to poor library construction (low-depth, high-sparsity, low library size) as technical errors. Patients were additionally excluded as outliers for meeting TAV-N criteria by aortic size but not being donor controls; these patients had undergone ascending aortic replacement at surgeon discretion with tissue collection during concomitant cardiac surgery.

Bioinformatics analysis was performed using established scripts in R. Differential gene expression was performed using the DESeq2 package in R^35^. Phenotype- and batch-independent quality control was performed using variance stabilization and established parameters for readability in RNA library construction (Supplemental Figure 1). Results of the differential gene expression were processed by log2 fold change (log2FC) and filtered by padj <0.1 for gene discovery. Gene IDs and symbols were matched to Ensembl IDs and ENTREZ IDs, respectively, using the org.Hs.eg.db database.

**Figure 1.**
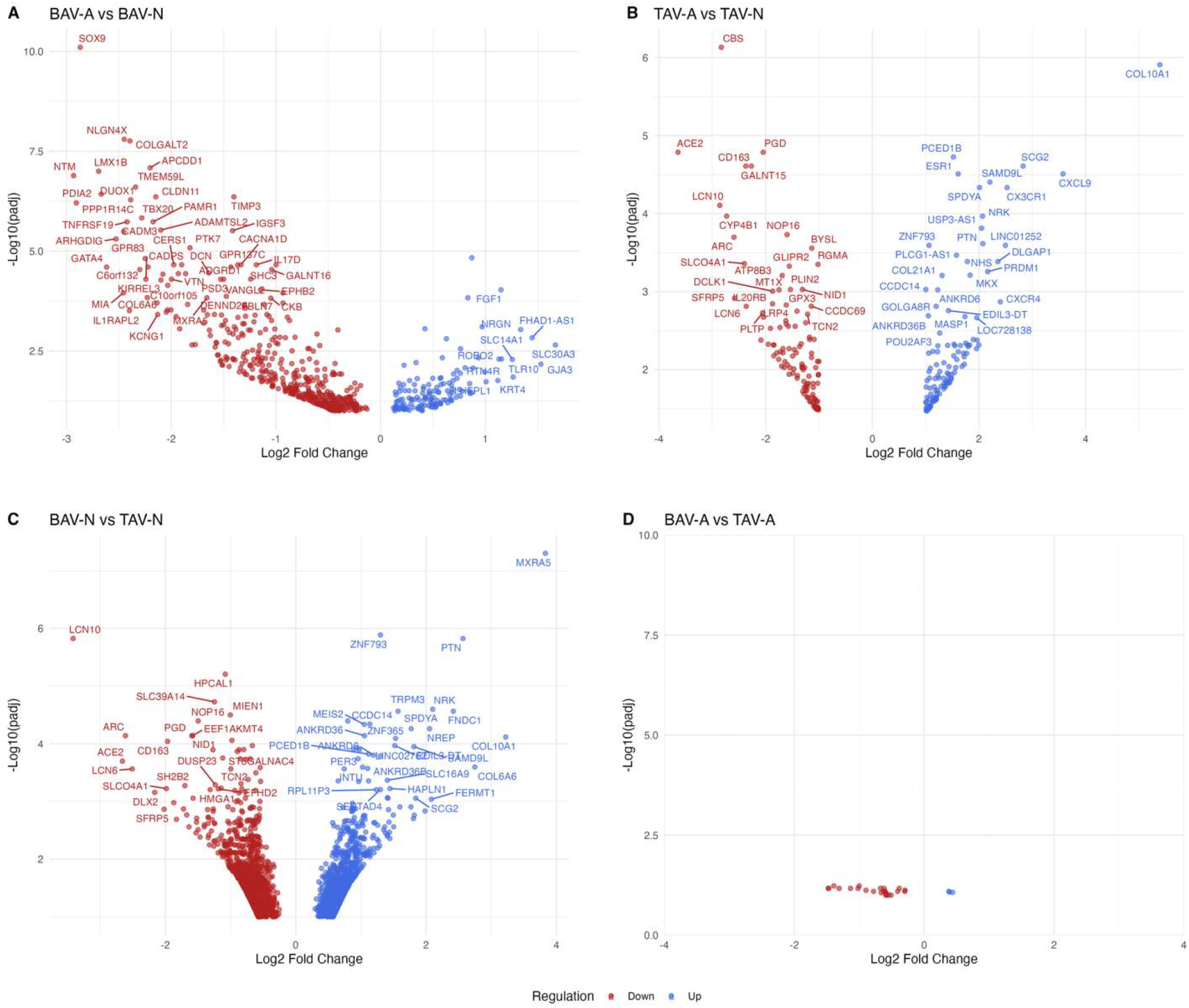
Differential gene expression by phenotype comparison: **(A)** BAV-A versus BAV-N, **(B)** BAV-N versus TAV-N, **(C)** TAV-A versus TAV-N, and **(D)** BAV-A versus TAV-A. Individual points represent genes that meet a filter of p-adj < 0.1. Labeled genes are those that meet a stricter filter of absolute log2 fold change > 1 and p-adj < 0.05.

Gene Ontology (GO) enrichment analysis was performed using the R package clusterProfiler and the reference database org.Hs.eg.db for Biological Process (BP), Molecular Function (MF), and Cellular Component (CC) for each phenotype^36^. A p-value cutoff of 0.1 and a q-value cutoff of 0.2 with the Benjamini-Hochberg p-adjust method were utilized.

Weighted Gene Co-expression Network Analysis (WGCNA) was performed in R on the differential gene expression dataset after filtering for genes with less than 15 counts in more than 75% of samples as suggested by the WGCNA for RNASeq protocol^37^. Variance stabilization was performed and batch effect removed prior to WGCNA. A soft threshold of 9 was selected as the smallest power meeting R^2^ ≥ 0.75 while preserving mean connectivity. Module eigengenes (MEs) were constructed using blockwise modules with this soft threshold, biweight midcorrelation (bicor), deepSplit = 4, minimum Module size of 25, and mergeCutHeight (merge threshold) of 0.10. Driver (hub) genes were identified using a composite driver score summing the correlation between each differentially expressed gene (DEG) and the ME (kME), intermodular connectivity of the DEG within the module (kWithin), and gene significance (GS) defined as the correlation between a DEG and the phenotype. Network mapping was visualized in Cytoscape^38^.

Cell-type specificity analysis of differentially expressed genes was determined using the R package scMappR and the established Aorta single cell RNA sequencing matrix “SRA734405_Aorta” within the package as the reference single-cell transcriptome^39^.Briefly, the filtered DEGs with padj < 0.1 were normalized, then the cell-type proportion for each DEG calculated in both the phenotype contrast and signature matrix resulting in an estimated cell-type proportion for each DEG. Cell-type specificity was then assigned to the fold-change of each DEG. Cell-weighted fold change (cwFC) of DEGs in the bulk RNA-seq sample were calculated as a function of fold-change, cell-type specificity, and cell-type proportions. A cwFC > 1 for a DEG means that a specific cell-type contributes to the observed upregulation of that DEG, and a cwFC < 1 means that a specific cell-type contributes to the observed downregulation of that DEG. The greater the cwFC, the stronger the contribution. A cwFC = 0 means the cell-type does not contribute to the observed expression for a specific DEG.

## Results

Ascending aortic tissue samples from 60 patients undergoing cardiac surgery at the University of Utah were included in our analysis: 4 TAV-N, 16 BAV-N, 28 BAV-A, and 12 TAV-A. Demographic information is included in Table 1. Demographic information and pre-operative imaging were not accessible for TAV-N patients as they were all heart transplant donors. Average age was similar between the two BAV groups. TAV-A patients were on average approximately 10 years older than BAV patients, although this was not statistically significant. Most patients were male. BSA was similar across phenotypes. Only one patient in BAV-N (6.3%) and BAV-A (3.6%) had a family history of aortic disease compared to five patients (45.5%) in TAV-A. 90.9% of TAV-A patients had hypertension compared to 43.8% and 64.3% of patients with BAV-N and BAV-A, respectively. Average absolute aortic diameter was 3.8 ± 0.35 cm for BAV-N, 5.1 ± 0.38 cm for BAV-A, and 5.3 ± 0.44 cm for TAV-A, as measured on pre-operative CT Chest. 87.5% of patients with BAV-N had greater than or equal to moderate aortic stenosis (AS) at the time of surgery—most commonly their operative indication—versus 35.7% of BAV-A patients. None of the TAV-A patients had aortic stenosis.

**Table 1.** Demographic characteristics of the four phenotypes. P-value determined using ANOVA single-factor test except for average absolute aortic size in which a Welch’s t-test was used comparing (a) BAV-N and BAV-A, (b) BAV-A and TAV-A, and (c) BAV-N and TAV-N.

| Demographic Characteristic | Phenotype |  |  |  | p-value |
| --- | --- | --- | --- | --- | --- |
|  | TAV-N (n = 4) | BAV-N (n = 16) | BAV-A (n = 28) | TAV-A (n = 12) |  |
| Age (years) | n/a | $51 \pm 15.1$ | $54.9 \pm 11.3$ | $62.5 \pm 12.9$ | 0.07 |
| Male | n/a | 8 (50%) | 18 (64.3%) | 8 (72.7%) | 0.59 |
| BSA | n/a | $2.0 \pm 0.24$ | $2.1 \pm 0.37$ | $2.2 \pm 0.43$ | 0.50 |
| Family history of aortic disease | n/a | 1 (6.3%) | 1 (3.6%) | 5 (41.7%) | 0.002 |
| Hypertension | n/a | 7 (43.8%) | 18 (64.3%) | 10 (90.9%) | 0.10 |
| Average absolute aortic size (cm) | n/a | $3.8 \pm 0.35^{a,c}$ | $5.1 \pm 0.38^{a,b}$ | $5.3 \pm 0.44^{b,c}$ | $^{a} 2.08 \times 10^{-12}$<br>$^{b} 0.27$<br>$^{c} 6.0 \times 10^{-9}$ |
| Moderate+ aortic stenosis | n/a | 14 (87.5%) | 10 (35.7%) | 0 (0%) | $1.12 \times 10^{-6}$ |

Volcano plots of differentially expressed genes (DEGs) for the various comparisons are shown in Figure 1. There were a total of 590 unique DEGs in BAV-A versus BAV-N, 2,589 DEGs in BAV-N versus TAV-N, and 1,242 DEGs in TAV-A versus TAV-N with an adjusted p-value (p-adj) < 0.1 (Figure 1 A-C). Labeled genes in Figure 1 meet a stricter filter of absolute log2 fold change > 1 and p-adj < 0.05.

To understand the broader gene functions, cellular locations, and biological roles genes involved in BAV and TAV aortopathy, we used Gene Ontology (GO) enrichment analyses. GO analyses revealed differential enrichment of genes across each category—Biological Process, Molecular Function, and Cellular Component—between phenotype comparisons. GO enrichment within Biological Processes demonstrated an enrichment of DEGs associated with extracellular structure and extracellular matrix (ECM) organization in BAV-A vs BAV-N as well as TAV-A vs TAV-N, and regulation of leukocyte mediated immunity in BAV-N vs TAV-N (Figure 2A). These themes were consistent in Molecular Function enrichment (Figure 2B) and Cellular Component enrichment (Figure 2C) for both aneurysm contrasts. BAV-A vs BAV-N was additionally uniquely enriched in the MF Wnt-protein binding. Interestingly, while the GO enrichment categories for BAV and TAV aneurysms largely correlated to the same pathways, the individual DEGs driving those shared enrichment pathways were mostly different (Figure 2D-G). For example, down-regulation of *COL3A1, COL5A1, COL15A1, COL6A3*, and *COL9A3* specifically drive enrichment in ECM and collagen-related enrichments in BAV-A vs BAV-N, while up-regulation of *COL10A1, COL21A1, COL22A1* and down-regulation of *COL5A3* and *COL7A1* specifically drive those enrichments in TAV-A vs TAV-N. Looking at these shared enrichments pathways, BAV-A vs BAV-N features down-regulation of structural collagens, collagen processing proteins, integrins, and other ECM proteins suggesting a role for matrix disorganization in aortic wall assembly and loss of aortic elasticity. In contrast, TAV-A vs TAV-N features differential expression of genes related to vascular wall scarring and stiffness as well as degradation and response to inflammation within these ECM-related enrichment pathways. Across all three enrichment categories, BAV-N vs TAV-N differed from the aneurysmal contrasts with top processes being regulation of leukocyte mediated immunity, transcription co-regulator bind, and cell leading edge (Figure 2A-C).

**Figure 2.**
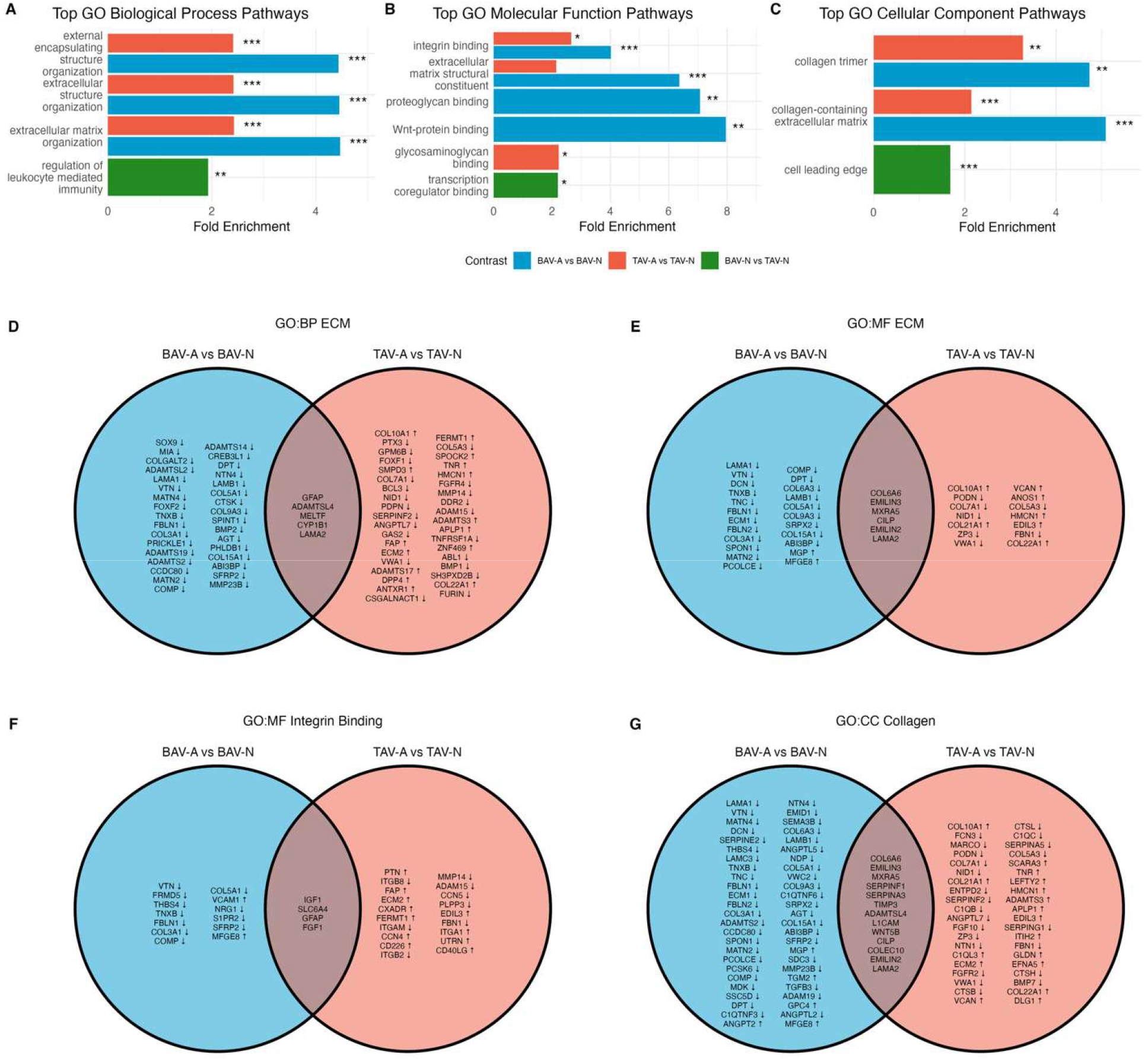
Enrichment analyses for each phenotype comparison. Genes comprising each GO sub-category between phenotypes for each GO category—**(A)** Biological Process, **(B)** Molecular Function, and **(C)** Cellular Component. Blue, BAV-A vs BAV-N; green, BAV-N vs TAV-N; orange, TAV-A vs TAV-N. Venn diagrams of genes comprising shared GO enrichment pathways in BAV-A vs BAV-N and TAV-A vs TAV-N; genes are ordered by abs(log2 fold change) greatest to least and up/down arrows represent up-/down-regulation within that phenotype comparison: **(D)** Biological Process – ECM, **(E)** Molecular Function – ECM, **(F)** Molecular Function – Integrin Binding, **(G)** Cellular Component – Collagen.

To identify transcriptional programs associated with valve and aneurysm phenotypes, we performed weighted gene co-expression network analysis (WGCNA). WGCNA is an unsupervised systems-level method that groups genes into modules based on shared expression patterns across samples^37^. DEGs were clustered into modules based on weighted expression correlations under the assumption that highly co-expressed genes participate in shared biological programs. Each module is identified by an eigengene representing the predominant expression pattern within the module, as determined by first principal component of all genes in the module. For clarity, we labeled the modules based on the function of the hub genes comprising each module; these are phenotype-agnostic. The module-trait correlation heatmap illustrates the relationships between module eigengenes (MEs) and valve-aorta phenotypes (Figure 3A). Across the groups, TAV-N samples demonstrated the strongest and most significant module-trait correlations, indicating highly coordinated transcriptional organization. For example, modules associated with endothelial integrity, ECM organization and homeostasis, signal transduction and adhesion, immune cell activation and signaling, intracellular functions, and cellular homeostasis were positively correlated in TAV-N samples, while cellular differentiation, vascular cell structure, and protein translation and trafficking modules were inversely correlated. BAV-N samples also demonstrated a positive correlation with the ECM organization and homeostasis module and inverse correlation with the vascular cell structure module. In contrast, the aneurysmal samples (BAV-A and TAV-A) showed weaker module-trait associations overall. As well, both aneurysmal phenotypes demonstrated negative correlation with the ECM organization and homeostasis module and positive correlation with the vascular cell structure modeule—the reverse of what is observed in the non-aneurysmal phenotypes. Taken together, these findings suggest that a normal aorta in the context of a trileaflet aortic valve maintains tightly coordinated transcriptional programs with an observed deviation but still relative coordination in the context of a bicuspid valve, whereas the pathological aneurysmal states are characterized by greater deviation or attenuation of these coordinated programs.

**Figure 3.**
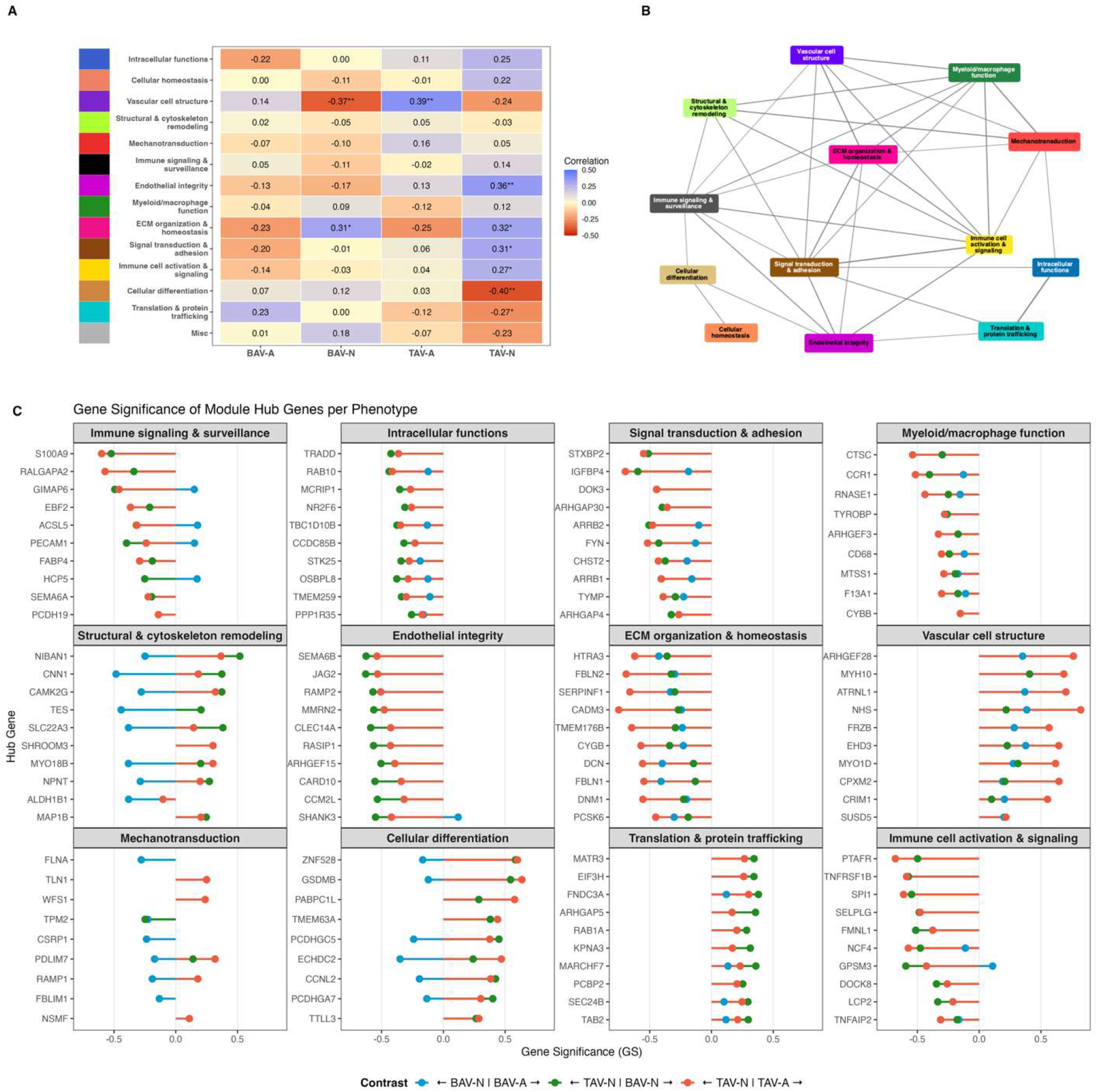
**(A)** Module eigengene (ME)-phenotype correlation heatmap where positive correlation (blue) represents upregulation of that module in the phenotype and negative correlation (red) represents downregulation; MEs are delineated by color and defined based on dominant hub genes’ functions. **(B)** Network map, where nodes are MEs and edges represent interconnectivity. **(C)** Gene significance (GS) representing the correlation between gene expression and phenotype for top GS hub genes per top 12 modules per phenotype comparison BAV-A vs BAV-N (blue), BAV-N vs TAV-N (green), and TAV-A vs TAV-N (orange); negative GS represents the gene is associated with the latter phenotype, positive GS represents the gene is associated with the former phenotype, and GS = 0 means no association between gene and either phenotype; magnitude of GS represents strength of gene association with phenotype.

The interconnectivity of the module eigengenes is illustrated in Figure 3B, in which nodes represent module eigengenes and the edges between nodes denote the strength of the correlation between modules. The ECM organization and homeostasis module occupied a central position within the network, with several connections to other modules including vascular cell structure, signal transduction and adhesion, and mechanotransduction. This indicates ECM organization and homeostasis, which relates to aortic elasticity, is tightly coordinated with other transcriptional programs in aortic tissue. Signal transduction and adhesion as well as immune cell activation and signaling MEs also demonstrated strong interconnectivity within the network.

To determine if module eigengenes significantly correlated with phenotype were driven by biologically relevant genes, we examined gene significance of hub genes within the modules (Figure 3C; Supplemental Figure 2). Gene significance (GS) represents a calculated correlation between gene expression and phenotype. Within a phenotype comparison, positive GS favors the former phenotype, negative GS favors the latter phenotype, and GS = 0 means no association between gene expression and either phenotype. Hub genes demonstrated consistent directionality and magnitude of association across phenotype contrasts, indicating that the most highly connected genes within each module are also strongly linked to valve-aorta phenotype. These findings reinforce the module-trait relationships observed in Figure 3A and the structured inter-module connectivity shown in Figure 3B, supporting the conclusion that coordinated, network-level transcriptional programs across a plethora of different categories distinguish normal and pathological aortic states.

Next, we sought to infer which cell types preferentially contribute to the phenotype-specific transcriptional changes. DEGs from each phenotype contrast were projected onto an aortic single-cell transcriptome reference matrix to compute cell-weighted fold changes^39^. Briefly, deconvolution tools were used to infer the relative cell-type proportions of a bulk RNA-seq sample based on a single-cell RNA-seq reference, generating a cell-weighted fold change (cwFC) as the proportion of each DEG fold change per cell-type specific expression of that gene. This analysis identified aorta cell type-specific signatures that are consistent with the observed bulk transcriptional differences as well as revealed cell type-specific contributions to each contrast (Figure 4). For bicuspid aortic valves, differential gene expression in the aneurysmal phenotype associated with the ECM, cell adhesion, and signaling appeared to be associated with vascular smooth muscle cells (VSMCs), mesenchymal cells, and matrix fibroblasts (Figure 4A). Differential gene expression minimally correlated with immune cells in the BAV-A phenotype relative to normal BAV aortas. Trileaflet aortic valve aneurysms similarly saw differential gene expression in ECM, adhesion, and signaling categories associated with VSMCs, mesenchymal cells, and matrix fibroblasts, although the genes were largely different than those corresponding to these categories in the BAV-A vs BAV-N phenotype comparison (Figure 4B). Unlike aneurysmal BAV aortas, the aneurysmal TAV phenotype suggested a stronger immune/inflammatory transcriptional component, underscored by up- and down-regulated genes mapping to T-cells, NK cells, myeloid cells, and macrophages. Between non-aneurysmal BAV and TAV aortas, a mix of up- and down-regulation of genes was associated with VSMCs, matrix fibroblasts, mesenchymal cells, and myeloid cells/macrophages, although the majority of DEGs did not cluster into defined GO categories (Figure 4C). Here, a role for other immune cells was not clearly defined. There were few transcriptional differences associated with distinct cell types suggested between aneurysmal states (BAV-A vs TAV-A; Figure 4D). Across all phenotype comparisons, differential gene expression did not map to endothelial cells. Altogether, these data suggest baseline structural and immune transcriptional differences between BAV and TAV aortas that disappear at the final aneurysmal phenotype. The unique transcriptional signatures of BAV and TAV aneurysmal and non-aneurysmal aortas in this context suggest different mechanisms of aneurysmal development based on baseline aortic differences corresponding to valve morphology.

**Figure 4.**
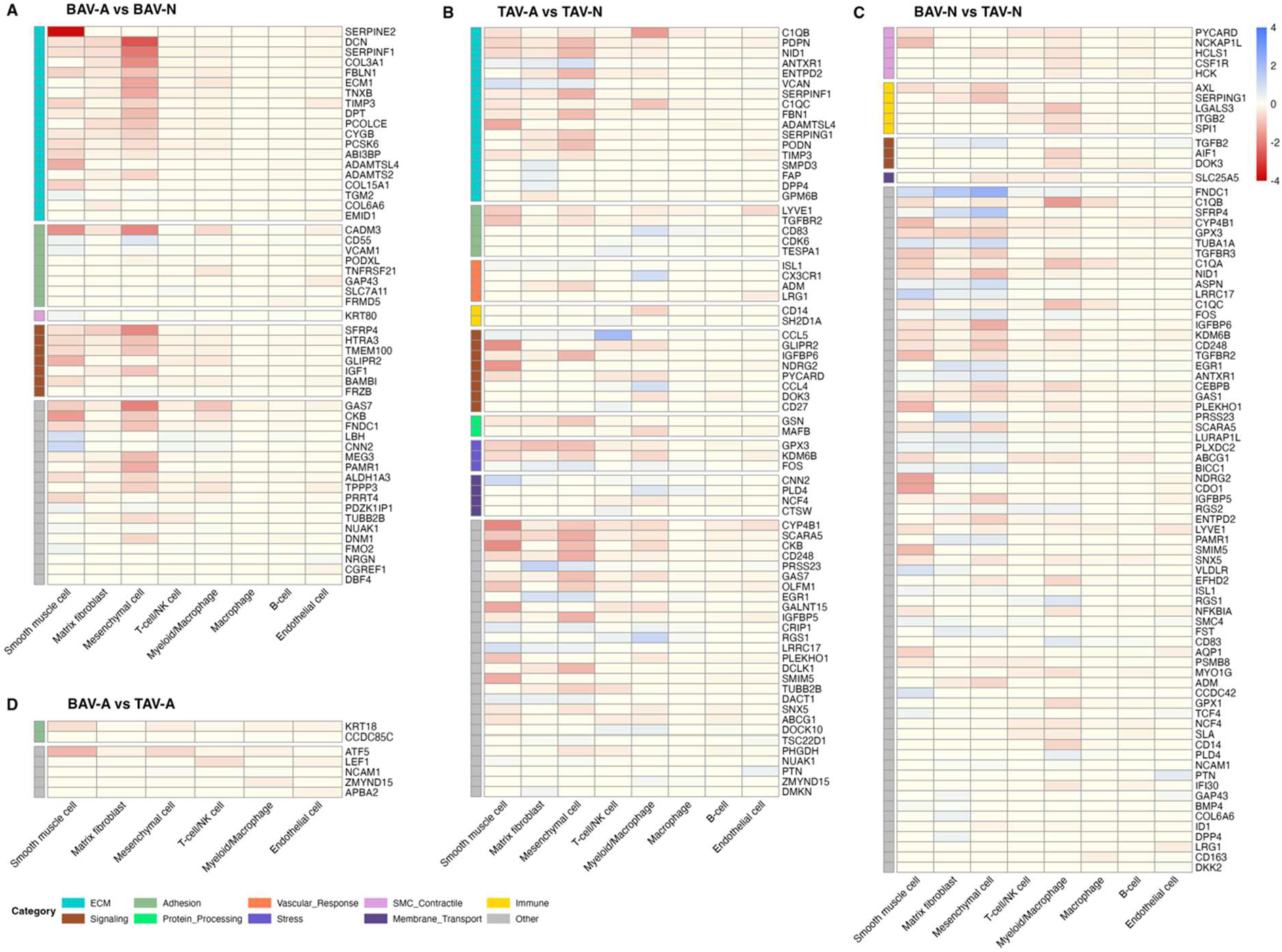
Projection of phenotype-associated cell-weighted fold changes onto a single-cell aortic signature matrix reveals cell type-specific contributions to each contrast: **(A)** BAV-A vs BAV-N, **(B)** TAV-A vs TAV-N, **(C)** BAV-N vs TAV-N, and **(D)** BAV-A vs TAV-A. Rows include DEGs driving the phenotype contrast; columns include the single cell-defined aortic cell types; heatmap red-blue color gradient represents the scaled cell-weighted fold-change; the lefthand margin colored markers represent category based of composite GO BP, MF, and CC categories.

## Discussion

BAV aortopathy is a complex disease process influenced by both genetics and the environment. Despite multiple attempts to elucidate these mechanisms, much is still unknown. Unlike heritable syndromic aortopathies, BAV aortopathy is likely polygenic with incomplete penetrance. Our study is the first robust transcriptomics analysis of BAV aortopathy in humans to date. We hypothesized that BAV aortopathy progresses through a distinct molecular mechanism as compared to trileaflet aortic valve aneurysms. Of the 60 patients included in our analysis, 16 were non-aneurysmal BAV and 28 were aneurysmal BAV. This afforded a strong cohort to test our hypothesis.

First, we demonstrated differentially expressed genes (DEGs) in non-aneurysmal aortic tissue as well as in aneurysmal tissue. DEGs were filtered liberally to maximize potential hits while minimizing the false discovery rate. In comparison of aneurysmal versus non-aneurysmal BAV aortas, there were preferentially more down-regulated than up-regulated DEGs. In non-aneurysmal BAV versus TAV aortas, there was a more equivalent mix of up- and down-regulated DEGs including statistically significant genes. This pattern of differential gene expression was similarly observed in aneurysmal versus non-aneurysmal TAV aortas. However, there were minimal DEGs in comparison of aneurysmal BAV and TAV aortas with no statistically significant genes. Altogether, this provides evidence that baseline BAV aortic tissue is transcriptionally distinct from TAV aortas. In contrast, there appears to be a shared aneurysmal transcriptome between BAV and TAV aortas. While there were more than 20,000 DEGs identified in comparison of aneurysmal BAV versus TAV aortas, the differences were so minute such that no DEGs met a significance threshold of absolute log2 fold change > 1 and an adjusted p-value (p-adj) < 0.05. Looking at the raw differential gene expression results for BAV-A versus TAV-A, only seven DEGs meet a threshold of log2 fold change < -1 (there were zero DEGs meeting a threshold > 1), and these DEGs have non-significant p-adj values of 0.059-0.068. The minimal differences in filtered DEGs in the aneurysmal comparison suggests that the differences in gene expression are not biologically significant and that the two phenotypes have a high degree of transcriptional similarity. This supports the hypothesis that BAV and TAV aortopathies develop through distinct mechanisms related to their baseline biological differences but converge to a common final transcriptomic pattern.

Aneurysmal vs non-aneurysmal BAV and TAV aortas demonstrated shared enrichment in gene ontology (GO) pathways related to extracellular matrix structure and organization. However, the involved genes largely differed for the two phenotype comparisons. For example, 33 down-regulated genes and 37 up/down-regulated genes mapped to GO-Biological Process (BP) extracellular matrix (ECM) pathways for BAV-N and TAV-N, respectively; only five genes were shared between BAV-N and TAV-N in GO-BP enrichment of ECM pathways. This pattern was also observed in the GO-Molecular Function (MF) ECM, GO-MF integrin binding, and GO-Cellular Component (CC) collagen trimer/collagen-containing ECM pathways. Together, this suggests that while there are shared pathways implicated in aneurysmal development for BAV and TAV aortas, the actual genes driving the change are distinct between the two valve phenotypes. We observed several transcriptional changes that suggest cell-matrix disorganization including the down-regulation of *COL3A1* implicated in vascular Ehlers-Danlos^40^, *COL5A1* regulating fibrillogenesis and implicated in aortic dissection^41^, *TNXB* (tenacin XB) regulating elastic fiber structure and stability^42^, several laminin genes, *VTN* encoding the ECM glycoprotein vitronectin implicated in cardiovascular hemostasis^43^, and *FBLN-1* encoding the ECM protein fibulin-1 highly expressed in the aortic media and associated with aortic wall stiffness^44^. This suggests a mechanism to explain the loss of aortic elasticity that may underlie aneurysm development in BAV aortas.

In weighted gene co-expression network analysis (WGCNA), co-expression of genes related to structural and cytoskeleton remodeling and ECM organization and homeostasis including *CNN1, DCN, FBLN1*, and *FBLN2* is more conserved in non-aneurysmal BAV aortas compared to aneurysmal BAV aortas (negative gene significance observed). *CNN1* encodes the actin filament-associated regulatory protein calponin-1, a key contractile protein in smooth muscle cells^45^. The proteoglycan decorin (*DCN*) aids in the spatial organization of the aortic elastic matrix and has been shown to be highly expressed in fibroblasts from TAAs^46,47^. Decreased gene and protein expression of fibulin-1 has been previously observed in acutely dissected aortas^48^, and the role of fibulin-2 (*FBLN2*) in the differentiation of vascular smooth muscle cells (VSMCs) from mesenchymal stem cells (MSCs) and ECM-SMC organization of the aorta during development has been described^49^. As well, overall, our co-expression analysis identified key modules with genes associated with “abnormal” phenotypes—non-aneurysmal BAV, aneurysmal BAV, and aneurysmal TAV. These modules and their hub genes are potential targets for further genomic studies for clinically relevant pathogenic variants.

Aneurysmal vs non-aneurysmal BAV aortas also demonstrated a significant enrichment in genes related to Wnt protein binding not observed in TAV aortopathy. The Wnt signaling pathway, a central cellular differentiation pathway for many cell types whose dysregulation has been implicated in aneurysm development, specifically correlated with BAV aortopathy in our data. Kostina et al demonstrated activation of the Wnt/β-catenin signaling pathway in aneurysmal thoracic aortic endothelial cells attenuates endothelial integrity, and these cells have a decreased response to shear stress resistance via the Wnt signaling pathway^50^. Their group also previously demonstrated that both BAV and TAV aneurysms demonstrate decreased VSMC and endothelial cell proliferation rates with increased collagen I and decreased fibrillin and elastin content in the aortic media compared to heart transplant donor controls^51^. Together, decreased proliferation, reduced response to stress, and abnormal ECM composition may underlie aneurysm biology. In our investigation, we observed suspected changes in VSMC-related genes but not significantly in endothelial-related genes.

Aortic tissue is heterogeneous in cell composition across the three aortic layers, and this granularity cannot be defined with bulk RNA sequencing. However, deconvolution cell-weighted simulated single-cell level investigation scMapp analysis suggests that BAV aortopathy is driven by matrix fibroblasts, VSMCs, and MSCs. These cells are found in the aortic media and adventitia—layers that are also rich in ECM components such as elastin, collagen, fibrillin-1, fibulins, microfibrils, and proteoglycans. There is evidence that VSMCs in thoracic aortic aneurysms (TAAs) switch from a differentiated contractile phenotype to a dedifferentiated state characterized by increased cellular proliferation, reduced apoptosis, and increased expression of proteolytic enzymes for adaptive ECM remodeling^52^. Nataatmadja et al demonstrated increased VSMC apoptosis in the absence of inflammatory cells as well as abnormal ECM deposition in BAV and Marfan’s syndrome TAAs compared to normal trileaflet aortic valve TAAs, suggesting that an intrinsic cell defect in VSMCs underlies these pathologies as opposed to an inflammatory state ^53^. Our data suggest that BAV-A tissue has a decreased contractile transcriptome, supporting the mechanistic hypothesis that VSMC dedifferentiation is associated with BAV aortopathy.

Cell-specificity analysis further demonstrates consistencies with the literature. We found that *SERPINE2* expression was notably downregulated in a pattern consistent with VSMCs in BAV-A vs BAV-N. *SERPINE2* encodes the glycoprotein serpinE2 which acts as a serine protease inhibitor; its expression has been observed in VSMCs, fibroblasts, and endothelial cells and been showed to have a role in development of cardiac fibrosis^54^. Knockdown of *SERPINE2* in cardiac fibroblasts has been shown to result in decreased extracellular collagen content in a cardiac fibrosis heart failure model^54^. Our BAV-A vs BAV-N cohort also demonstrated a downregulation of *ADAMTS2* and *ADAMTSL4* expression corresponding to smooth muscle cells. ADAMTS—a disintegrin and metalloproteinase with thrombospondin motifs—are integral to the turnover of ECM proteins in vascular tissues^55^. ADAMTS-like (ADAMTSL) proteins are part of the ADAMTS superfamily and interact with specific extracellular binding proteins but lack the catalytic domain of ADAMTS proteins. Knockdown of various *ADAMTS*s and *ADAMTSL*s in rodents have demonstrated aortic dilation with abnormal ECM content^55–58^, although our observed downregulation of *ADAMTSL4* and *ADAMTS2* has not been previously reported.

These data also highlight differential gene expression in mesenchymal cells but not in endothelial cells in aneurysmal BAV aortas compared to non-aneurysmal BAV and both TAV aorta phenotypes. One hypothesis generated here is that aneurysmal BAV aortas have an increased endothelial-to-mesenchymal transition (EndMT) of cells within the aortic wall. External factors such as stress and epigenetic changes activate TGF-β, WNT/β-catenin, and NOTCH signaling pathways, which trigger endothelial cells to acquire mesenchymal cell features^59^. This results in vascular tissue with junction instability and increased permeability. As well, we observed differential cell-weighted expression of genes in mesenchymal cells in across phenotype comparisons except for BAV-A vs TAV-A. The most notable differences were observed with BAV-A vs BAV-N with down-regulation of *DCN, SFRP4, SERPINF1*, and *COL3A1*. Secreted frizzled-related protein 4 (SFRP4) and SERPINF1 have been shown to inhibit angiogenesis^60,61^.

Conversely, the differential gene expression pattern in aneurysmal vs non-aneurysmal TAV aortas points towards a distinct mechanism for aneurysmal development from BAV aortopathy. TAV-A vs TAV-N specifically demonstrated differential expression of genes related to vascular response to injury and fibrosis—*PTX3* (pentraxin-3)^62^ and *FAP* (fibroblast activation protein)^63,64^—as well as the FACIT collagens *COL21A1* and *COL22A1* in the ECM implicated in maintaining vascular stability^65,66^. In co-expression analysis (WGCNA), non-aneurysmal TAV aortas demonstrated highly correlated modules eigengenes across a plethora of categories, indicating a tightly coordinated transcriptional program in normal aortic tissue. The attenuation of this tight control was observed in aneurysmal TAV in cellular homeostasis, ECM organization and homeostasis, and immune signaling and surveillance modules. Interestingly, there was greater correlation between vascular cell structure gene expression observed in TAV-A compared to TAV-N. Altogether, this suggests two potential contributing mechanisms to TAV aortic aneurysmal development: dysfunction of an intrinsic cellular and ECM remodeling response to stress and an immune system role in maintaining aortic tissue homeostasis and integrity. The active role of T-cells, myeloid cells, and macrophages was demonstrated in our single-cell simulation. The observed differential expression of several genes in aneurysmal vs non-aneurysmal TAV aortas mapped to these immune cells; notable was the up-regulation of cytokine-encoding genes *CCL4* and *CCL5* in macrophages and T-cells, respectively. Increased active immune cell populations in thoracic aortic aneurysm tissue relative to non-aneurysmal aortic tissue has been described in a single cell-RNA seq analysis of a small mixed BAV/TAV aortic aneurysm cohort^47^.

Interestingly, there were no statistically significant DEGs between BAV and TAV aneurysms, suggesting a convergence of transcriptional phenotype at the final aneurysmal state. This is further supported by the minimal differences observed in our cell-weighted analysis. One explanation for these observations is that aneurysm development in BAV and TAV aortas occurs through different mechanisms related to their non-aneurysmal baseline states, as demonstrated by the observed differences in BAV-A vs BAV-N and TAV-A vs TAV-N, but once aneurysm development reaches a late stage, the biological differences in these tissues recedes and a final common phenotype is observed (i.e. they are transcriptionally indistinguishable).

There are several limitations to this study. First, we utilized bulk-RNA sequencing. While this method affords a global picture of differential gene expression, aortic tissue is heterogenous, and we cannot specifically delineate differential gene expression by cell type. The scMappR package allows us to extrapolate single cell-level interpretations, although these are limited and biased by the algorithm and preset reference signature matrices. Second, we have potential sampling bias in our cohort. All study participants were enrolled from our cardiac surgical practice. This represents an extreme phenotype of patients who required surgery, which may not be generalizable to the larger population. As well, not all patients consent to research, and of those who consent, disease phenotype is unevenly distributed. Thus, our study captures a portion of the population who presents for surgery but may not be representative of the at-large population due to the enrollment process. Relatedly, there is an unexpected higher rate of family history of aortic disease in observed in our aneurysmal TAV patients compared to our BAV patients, despite the consensus that BAV aortopathy has a heritable component. This may be an additional sampling bias as patients with a family history of aortic aneurysm/dissection may be more likely to have screening for aortic disease and subsequent intervention, while BAV and BAV aortopathy are often clinically silent. Third, transcription is a dynamic process that reflects the cellular microenvironment influenced by external factors and epigenetics. We do not address the potential influence of hemodynamics from BAV, variation in thoracic aortic shape, aortic stenosis, aortic insufficiency^67–69^, and/or hypertension in our analysis. As postulated by others, hemodynamic factors may contribute but do not fully explain BAV aneurysm development in BAV. Further sub-analysis of our bulk RNA sequencing data, additional sequencing from different regions of the ascending aorta, and single-cell RNA sequencing are next steps that may elucidate the interplay between hemodynamics and genomic factors on aneurysm development.

## Conclusion

Our findings provide evidence for three conclusions. The transcriptomes of non-aneurysmal thoracic aortas in patients with bicuspid aortic valve and trileaflet aortic valve are distinct. Further, aneurysmal development in aortic tissues from bicuspid and trileaflet aortic valve patients demonstrate distinct gene expression, suggesting mechanistic differences in aneurysm development (aortopathy) between these two populations. Finally, the observed transcriptional pattern is similar between aneurysmal bicuspid and trileaflet aortas, suggesting a final common transcriptional state for thoracic aortic aneurysm. These findings will provide the foundation to interpret future whole genome sequencing and genome-wide association studies to elucidate the potential genome-level susceptibility of bicuspid aortic valve patients to developing aortopathy.

## Data Availability

The data including R code referred to in the manuscript can be available by written request to the corresponding author.

